# Genetic analysis of female genital tract polyps implicates genome stability, estrogen signalling and shared susceptibility with proliferative gynaecological disorders

**DOI:** 10.64898/2026.04.13.26350740

**Authors:** Nathan Ingold, Stephanie Frankcombe, Kelsie Bouttle, Emily Moro, Daffodil Canson, Sebastian Zöllner, Snehal Patil, Jelisaveta Džigurski, Dylan M. Glubb, Triin Laisk, Tracy A. O’Mara

## Abstract

Female genital tract (FGT) polyps are common benign growths affecting up to half of all women. However, they carry malignant potential, and their genetic architecture remains poorly defined. We conducted a genome-wide association study (GWAS) meta-analysis across four biobanks (48,400 cases, 477,134 controls), identifying 26 risk loci for FGT polyps, 12 of which were previously unreported. Integrative gene prioritisation highlighted 193 candidate genes, revealing a potential convergent biological mechanism: where germline variation in DNA replication and maintenance (e.g., *PRIM1*, *TERT* and *HMGA1*) compromises genomic stability in the context of hormone-driven proliferation (e.g., *ESR1* and *GREB1*). This susceptibility is further modulated by metabolic drivers of estrogen biosynthesis, underscored by specific adiposity-related loci (e.g. *RSPO3* and *PLCE1*) and the aromatase gene *CYP19A1*. Mendelian randomisation demonstrated bidirectional causal relationships with endometriosis and fibroids, and endometrial cancer. Leveraging the shared genetic architecture of FGT polyps and other gynaecological disorders via multi-trait analysis revealed an additional 26 loci, validating sub-threshold regions encompassing *HMGA1* and *GREB1.* In total, 52 risk loci were identified (36 novel), 39 of which replicated in an independent cohort. These findings reframe polyps not merely as local gynaecological overgrowths but as manifestations of a systemic proliferative syndrome characterised by dysregulated genome stability and estrogen signalling, which may also impact malignant transformation.

## Introduction

Female genital tract (FGT) polyps are benign lesions that can form on the lining of the uterus, cervix, vagina, or vulva. Among these, endometrial (uterine) polyps are the most common, affecting between 8% - 50% of women, depending on the population studied ^1–3^. Many women with FGT polyps experience abnormal uterine bleeding, a symptom reported in nearly two-thirds of diagnosed cases ^4^. Yet, estimates suggest that up to 80% of polyps remain asymptomatic - often evading detection ^1,2^. Although considered benign, the potential for malignancy of uterine polyps to form endometrial cancer ranges from 2.5% in women under 35, to as high as 37% in those over 65 years old ^5^.

The pathogenesis of FGT polyps remains unclear, as does an understanding of the progression from polyp to malignant neoplasm. Nonetheless, both estrogen and non-estrogen response-related pathways have been associated with polyp onset ^6^. Genetic risk variation may help further reveal underlying biological driver mechanisms. Currently, the largest genome-wide association study (GWAS) of FGT polyps identifies 16 genetic loci associated with FGT polyps risk, explaining approximately 3% of FGT polyp variance (on the liability scale) ^7^.

Here, we performed a GWAS meta-analysis of FGT polyp risk across four cohorts to identify genetic susceptibility loci. Fine-mapping, gene-based and transcriptomic analyses were performed to prioritise candidate causal variants and risk genes from GWAS. Shared genetic architecture with related gynaecological traits was evaluated and leveraged to enhance locus discovery, with independent replication performed in the All of Us cohort ^8^.

## Results

Overall study design is depicted in **Figure 1**. This study was conducted under ethical approval of QIMR Berghofer and complied with all relevant cohort-specific ethical regulations (**Supplementary notes**).

**Figure 1.**
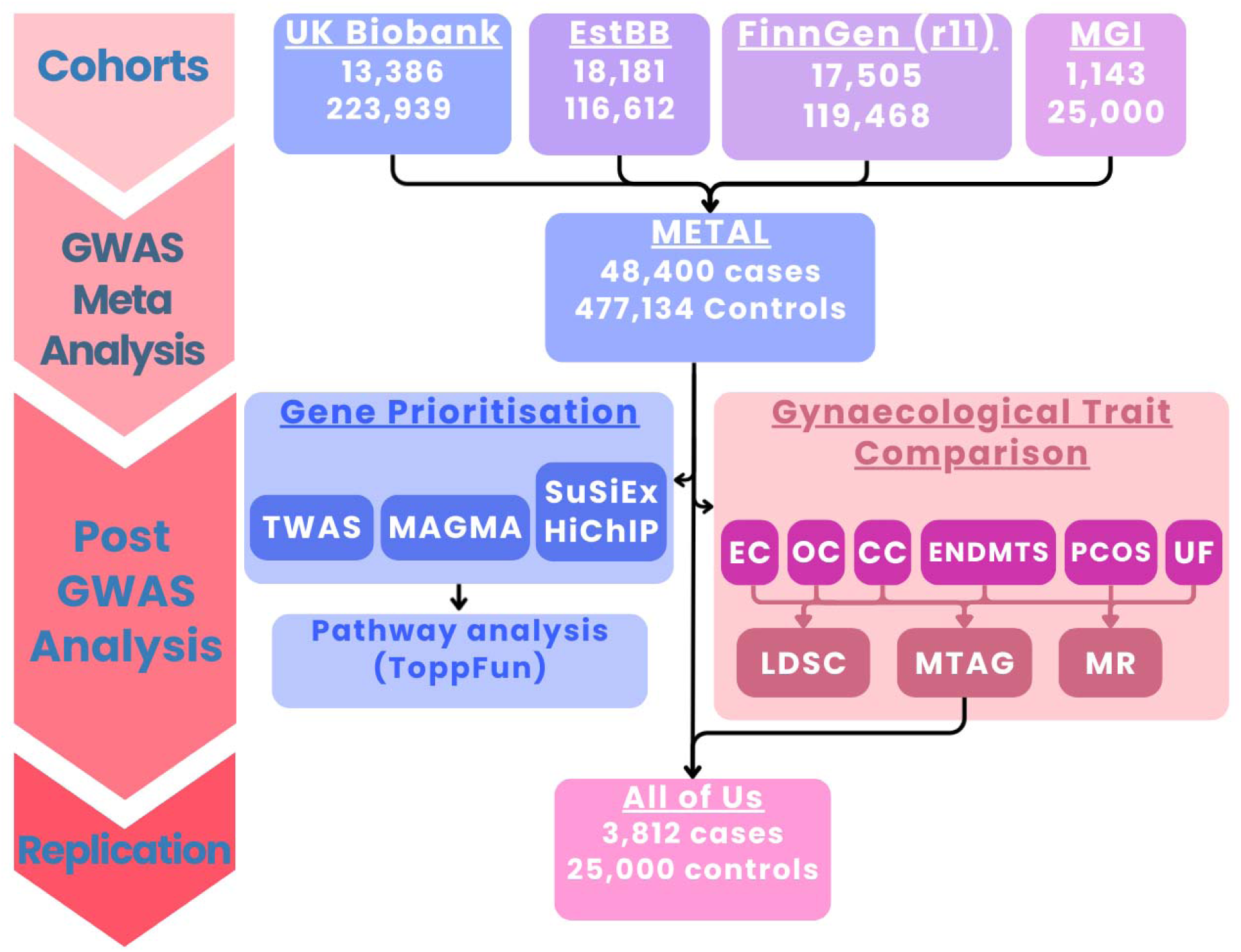
Study workflow and analytical pipeline. *The study design comprises three stages*: (1) Discovery: Meta-analysis of FGT polyp GWAS summary statistics from four independent cohorts (UK Biobank, Estonian Biobank, FinnGen, Michigan Genomics Initiative; total 48,400 cases, 477,134 controls). (2) Post-GWAS Analysis: The discovery meta-analysis informed two parallel downstream pipelines: Left: Gene prioritisation integrating TWAS, MAGMA, and fine-mapping (SuSiEx) intersected with H3K27Ac HiChIP chromatin contacts from endometrial epithelial cells, followed by functional enrichment (ToppFun). Right: Comparison with gynaecological traits (endometrial cancer [EC], ovarian cancer [OC], cervical cancer [CC], endometriosis [ENDMTS], polycystic ovarian syndrome [PCOS], uterine fibroids [UF]) using genetic correlation (LDSC), multi-trait analysis (MTAG), and Mendelian randomisation (MR). (3) Replication: Validation of meta-analysis and MTAG findings in the independent All of Us cohort (3,812 cases, 25,000 controls).

### Meta-analysis identifies 26 FGT polyp risk loci

GWAS meta-analysis of 48,400 cases and 477,134 controls identified 26 loci associated with FGT polyp risk at genome-wide significance (P < 5×10^-8^; **Table 1 & Figure 2**), 12 of which were previously unreported ^7,9^. Four novel loci map to regions associated with sex hormone-related traits and gynaecological disease (rs12526447, rs28490942, rs2800708, rs5977667), with adjacent genes including *ESR1* and *CYP19A1*, directly implicating estrogen signalling in FGT polyp aetiology ^10,11^. Pairwise genetic correlations among contributing GWAS ranged from 0.74 to 1.0 (**Supplementary Figure 1**); significant heterogeneity was observed in 3 of 26 lead SNPs but was non-significant after multiple testing correction (**Supplementary Table 1**). SNP-based heritability was h² = 0.048 (95% CIs = 0.036–0.060), consistent with the range across input GWASs (0–0.07).

**Figure 2.**
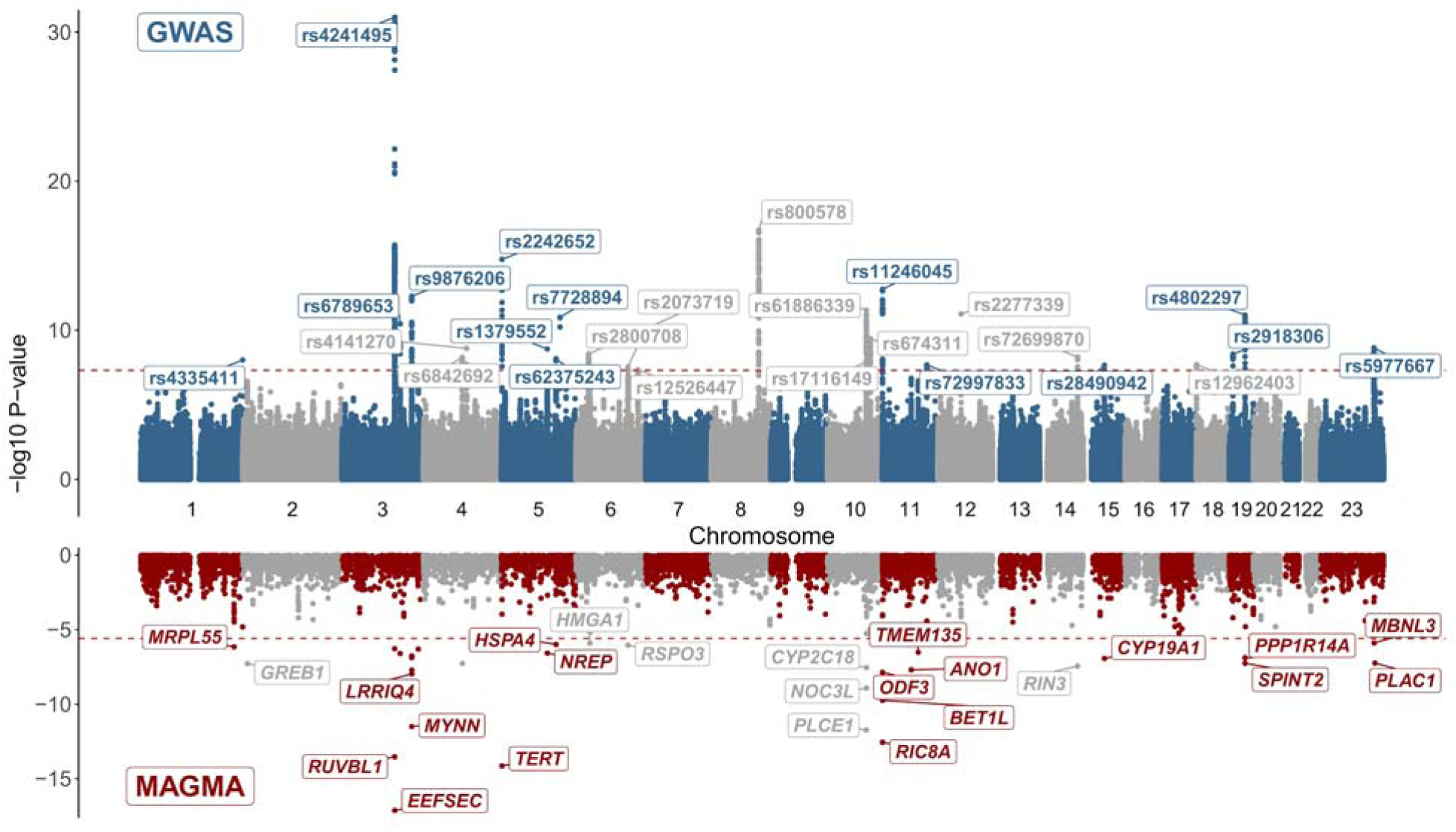
Genome-wide association meta-analysis and gene-based analysis of FGT polyps. Miami plot showing SNP-level (top) and gene-level (bottom) associations for FGT polyps (48,400 cases, 477,134 controls). Top: Manhattan plot of GWAS meta-analysis results; each dot represents one SNP. The 26 genome-wide significant loci (P < 5×10 ; red dashed line) are labelled with lead SNP rsIDs. Alternating blue and grey shading indicates chromosomes (1-23). P-values are displayed as −log (P), which have been capped at -log_10_(P) < 30 for visuali**s**ation purposes. Bottom: Gene-based association results from Multi-marker Analysis of GenoMic Annotation (*MAGMA) analysis; each data point represents one gene. Red labels indicate the 32 genes reaching genome-wide significance (P < 2.5×10 ; red dashed line). Only 25 of the 32 genes were labelled on the MAGMA plot (full list of significant genes in **Supplementary*** Figure 3 *and **Supplementary Table 4**)*.

**Table 1:** 26 lead SNPs from the GWAS meta-analysis of FGT polyp risk. *NB –* ◊ *marks a previously unreported locus, where a locus has multiple genes of interest, * highlights the nearest gene*.

| CHR | Cytoband | BP38 | SNP | A1* | A2 | A1FREQ | BETA | SE | P | Het PVal | Nearest Gene |
| --- | --- | --- | --- | --- | --- | --- | --- | --- | --- | --- | --- |
| 1 | q44 | 248897507 | rs4335411 | A | G | 0.75 | 0.05 | 0.009 | 1.0E-08 | 0.06 | PGBD2 |
| 3 | q21.3 | 128392331 | rs4241495 | T | C | 0.74 | -0.09 | 0.008 | 1.0E-31 | 0.98 | EEFSEC |
| 3 | q23 | 141432148 | rs6789653 | A | G | 0.34 | 0.05 | 0.007 | 3.6E-11 | 1.00 | ZBTB38 |
| 3 | q26.2 | 169799648 | rs9876206 | T | C | 0.26 | -0.06 | 0.008 | 5.1E-13 | 0.08 | ACTRT3/LRRC34*/MYNN |
| ◊4 | q22.3 | 94806355 | rs6842692 | T | G | 0.44 | -0.04 | 0.007 | 6.6E-09 | 0.01 | BMPR1B |
| ◊4 | q24 | 105101835 | rs4141270 | T | C | 0.12 | 0.06 | 0.011 | 1.6E-09 | 0.30 | TET2 |
| 5 | p15.33 | 1279913 | rs2242652 | A | G | 0.21 | 0.07 | 0.008 | 1.7E-15 | 0.43 | TERT |
| ◊5 | q22.1 | 111643461 | rs1379552 | T | C | 0.78 | -0.05 | 0.008 | 1.8E-09 | 0.08 | NREP/STARD4-AS1* |
| 5 | q31.1 | 133097605 | rs62375243 | A | G | 0.27 | -0.06 | 0.010 | 8.1E-09 | 0.32 | HSPA4 |
| 5 | q31.3 | 142783068 | rs7728894 | A | G | 0.27 | 0.05 | 0.008 | 1.4E-11 | 0.62 | ARHGAP26 |
| ◊6 | p21.33 | 31145148 | rs2073719 | T | C | 0.73 | -0.05 | 0.008 | 3.9E-09 | 0.83 | CCHCR1 |
| ◊6 | q22.33 | 127116472 | rs2800708 | T | C | 0.53 | 0.04 | 0.007 | 2.9E-08 | 0.06 | RSPO3 |
| ◊6 | q25.1 | 151718990 | rs12526447 | A | G | 0.70 | -0.04 | 0.007 | 4.4E-08 | 0.02 | ESR1 |
| 8 | q23.3 | 115857251 | rs800578 | T | C | 0.23 | 0.07 | 0.008 | 1.8E-17 | 0.06 | LINC00536*/TRPS1 |
| 10 | q23.33 | 94273109 | rs61886339 | T | C | 0.69 | -0.05 | 0.007 | 4.3E-12 | 0.13 | PLCE1 |
| 10 | q24.33 | 103928292 | rs17116149 | A | G | 0.04 | 0.11 | 0.020 | 1.1E-08 | 0.01 | NEURL1/OBFC1 |
| ◊10 | q25.1 | 104084134 | rs674311 | A | G | 0.96 | -0.15 | 0.024 | 3.7E-10 | 0.60 | STN1/COL17A1 |
| 11 | p15.5 | 274780 | rs11246045 | T | C | 0.90 | 0.09 | 0.012 | 1.7E-13 | 0.09 | BET1L/NLRP6*/ODF3 |
| ◊11 | q22.3 | 108492835 | rs72997833 | A | G | 0.18 | -0.05 | 0.009 | 2.1E-08 | 0.14 | ATM/POGLUT3* |
| 12 | q13.3 | 56752285 | rs2277339 | T | G | 0.89 | 0.08 | 0.011 | 8.2E-12 | 0.61 | PRIM1 |
| ◊14 | q32.12 | 92650006 | rs72699870 | T | C | 0.83 | -0.05 | 0.009 | 6.3E-09 | 0.13 | RIN3 |
| 15 | q21.2 | 51267648 | rs28490942 | C | G | 0.47 | 0.04 | 0.007 | 2.2E-08 | 0.05 | CYP19A1 |
| ◊18 | p11.32 | 672035 | rs12962403 | T | C | 0.15 | -0.05 | 0.010 | 2.0E-08 | 0.70 | TYMS |
| ◊19 | p13.2 | 8678357 | rs2918306 | T | C | 0.82 | -0.05 | 0.009 | 4.0E-09 | 0.25 | ACTL9 |
| 19 | q13.2 | 38247490 | rs4802297 | C | G | 0.54 | -0.05 | 0.007 | 9.8E-12 | 0.23 | PPP1R14A |
| ◊23 | q26.2 | 132262038 | rs5977667 | A | G | 0.30 | -0.06 | 0.009 | 1.5E-09 | 0.84 | RAP2C-AS1 |

Fine-mapping of lead loci with SuSiEx ^12^ identified 30 candidate causal variant (CCV) sets, comprising 634 CCVs among 24 (of 26) lead meta-analysis loci (**Supplementary Table 2**). Two loci, rs4335411 (1q44, near *PGBD2*) and rs2073719 (6p21.33, intronic to *CCHCR1*), could not be fine-mapped. The largest CCV set included 98 SNPs, while the smallest CCV set was 1 SNP. Seven CCVs fell in protein-coding sequences, including three synonymous variants in *MYNN* and *PLCE1*. Four were missense variants in *ODF3*, *PRIM1*, *LRRC34* and *ATM* (**Supplementary Table 3**), of which three (rs72878024 in *ODF3*, rs2277339 in *PRIM1* and rs10936600 in *LRRC34*) were predicted deleterious by SIFT or probably damaging protein effect (PolyPhen). Furthermore, rs2277339 had a posterior inclusion probability >0.8, providing strong statistical and functional evidence for *PRIM1* as a causal FGT polyp risk gene.

### Gene-based analysis reveals sub-threshold risk loci

To complement the GWAS meta-analysis and reduce the multiple testing burden, we performed Multi-marker Analysis of GenoMic Annotation (MAGMA) analysis. MAGMA identified 32 genes potentially associated with FGT polyp risk, representing loci tagged by aggregated SNP-level signals (q < 2.5×10^−6^; **Figure 2**, **Figure 3a; Supplementary Table 4**). Most genes were located at genome-wide significant GWAS loci. However, five genes (*MRPL55, TMEM135, ANO1, GREB1,* and *HMGA1)* had no corresponding genome-wide significant GWAS locus. Notably, *GREB1* is involved in estrogen receptor signalling, as is *HMGA1* ^13^ whose chromosomal rearrangements are detected in 74% of endometrial polyps ^14^. These genes may represent additional GWAS loci detectable with increased sample size.

**Figure 3:**
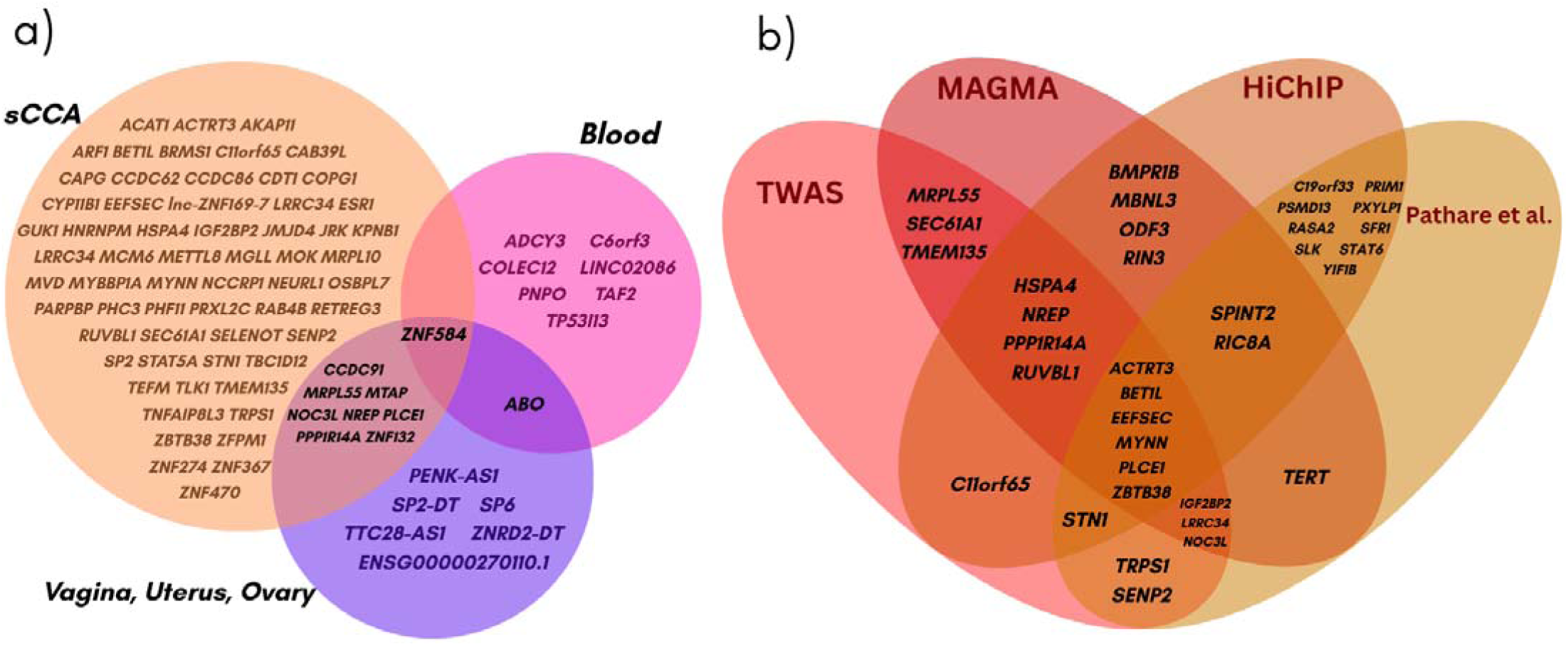
Prioritisation of FGT polyp candidate genes. a) Venn diagram showing the overlap of significant TWAS genes (FDR < 0.05) across three tissue categories: cross-tissue models (sCCA; brown), blood (whole blood and EBV-transformed lymphocytes; pink), and reproductive tissues (Uterus/Ovary/Vagina; purple). b) Venn diagram displaying the convergence of four distinct gene prioritisation strategies: TWAS, MAGMA (gene-based association), H3K27Ac HiChIP chromatin looping (linking GWAS variants to target gene promoters in endometrial cells), and previously reported candidate genes from Pathare et al. (2025). Genes in the overlapping regions are supported by at least two approaches. Supplementary Figure 2 and Supplementary Table 4 for the full list of prioritised genes.

### Functional mapping prioritises candidate genes

To identify candidate causal genes, we integrated transcriptome-wide association study (TWAS), MAGMA, and H3K27Ac HiChIP chromatin contact data. TWAS identified 85 genes whose predicted expression associated with FGT polyp risk, the majority detected using sparse canonical correlation analysis cross-tissue (sCCA) models ^15^ (**Figure 3a**). Fourteen genes showed tissue-specific associations in reproductive tissues or blood ^15^. Fifty-nine TWAS genes were not located at a GWAS locus, corresponding to 38 potential novel risk regions that may be revealed in larger studies (**Supplementary Table 5**); two of which (*MRPL55* and *TMEM135)* were also seen in MAGMA (**Supplementary Table 4**). HiChIP mapping linked FGT polyp CCVs within potential enhancers to 92 candidate target genes (corresponding to 634 fine-mapped CCVs + 2 lead GWAS meta-analysis SNPs that failed fine-mapping), 18 of which overlapped with TWAS and/or MAGMA findings (**Figure 3b**). Integrating all three approaches with candidates from Pathare et al. (2025)^7^ revealed 193 unique candidate genes (175 from our analyses plus 18 from Pathare et al.; **Supplementary Figure 2)**; 36 of which were supported by at least two prioritisation methods (**Figure 3b**).

### Pathway enrichment reveals biological mechanisms

Gene set enrichment analysis of the 193 candidate genes using ToppFun ^16^ revealed three dominant themes: DNA replication and genome maintenance, obesity-related traits, and estrogen signalling (**Supplementary Table 6**). A highly enriched biological process was DNA replication (q = 8.5 × 10^-3^), involving core replication machinery genes *PRIM1*, *CDT1* and *MCM6*. This was complemented by a strong enrichment for genes associated with telomere length maintenance (q = 2.95 × 10 ^6^), driven by *TERT,* alongside replication-telomere factor *STN1* and DNA damage response regulator *ATM.* We observed enrichment for genes associated with anthropometric traits such as body fat percentage (q = 2.09 × 10 ^3^) and waist-to-hip ratio (q = 5.79 × 10 ^3^), with candidate genes including *RSPO3*, *PLCE1* and *IGF2BP2*. Finally, there was significant enrichment in multiple gene sets related to estrogen signalling and hormone-dependent gynaecological disorders, encompassing key pathway regulators encoded by *ESR1* and *GREB1*, reinforcing the hormonal aetiology of polyp development.

### Genetic correlation with gynaecological disorders

We observed positive and significant genetic correlations between FGT polyps and endometrial cancer (r_g_ = 0.36, 95% confidence intervals [CIs] = 0.17 - 0.55), ovarian cancer (r_g_ = 0.28, 95% CIs = 0.1 - 0.45), endometriosis (r_g_ = 0.39, 95% CIs = 0.26 - 0.52), uterine fibroids (r_g_ = 0.39, 95% CIs = 0.28 - 0.51), PCOS (r_g_ = 0.2, 95% CIs = 0.02 - 0.37), but not cervical cancer (**Figure 4**; **Supplementary Table 7**). All correlations, except cervical cancer and PCOS, remained significant after Bonferroni correction for multiple testing.

**Figure 4.**
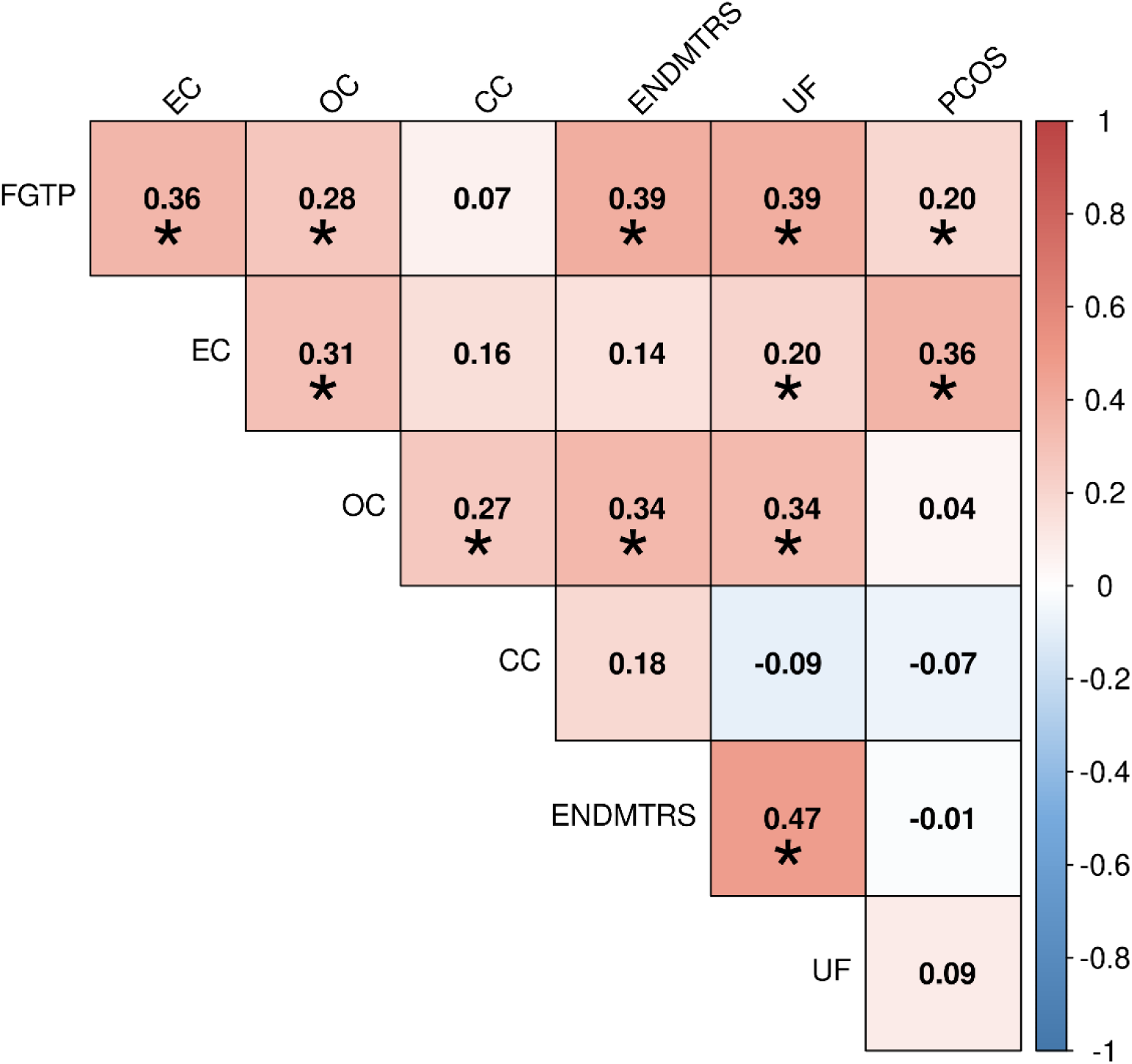
Genetic correlations between FGT Polyps and gynaecological disorders. Heatmap of pairwise genetic correlations (rg) estimated using LD score regression (LDSC) between FGT polyps (FGTP) and related gynaecological disorders: endometrial cancer (EC), ovarian cancer (OC), cervical cancer (CC), endometriosis (ENDMTRS), uterine fibroids (UF), and polycystic ovarian syndrome (PCOS). The colour scale represents the magnitude and direction of genetic correlation (red = positive, blue = negative). Asterisks (*) indicate significant correlations (P < 0.05). Bonferroni correction for multiple testing (P < 0.0083), see **Supplementary Table 7**.

### Bidirectional causal associations with gynaecological disorders

We performed Mendelian randomisation to assess causal relationships between FGT polyps and other gynaecological diseases and breast cancer ^17,18^. Bidirectional MR using 26 IVs (mean F = 45.5) identified significant causal associations between FGT polyp genetic liability and endometrial cancer (OR = 1.29, 95% CIs = 1.09–1.52), endometriosis (OR = 1.57, 1.36–1.82), and uterine fibroids (OR = 1.77, 1.46–2.14) (**Supplementary Figure 3 & Table 8**). All three relationships were significant in the reverse direction: genetic liability to endometrial cancer (OR = 1.15, 1.09–1.21), endometriosis (OR = 1.13, 1.05–1.21), and uterine fibroids (OR = 1.09, 1.04–1.15) each associated with increased FGT polyp risk (**Supplementary Figure 4 & Table 9**). Cervical and breast cancer showed unidirectional associations with polyp risk (OR = 1.07, 1.01–1.13; OR = 1.06, 1.02–1.09, respectively). Partial sample overlap was noted between several traits (**Supplementary Table 10**); however, sensitivity analysis using a non-overlapping endometrial cancer subset (which had the greatest sample overlap at 37.8%) ^19^ yielded a consistent effect (OR = 1.27, 95% CIs = 1.04–1.54).

### Multi-trait analysis reveals 26 additional risk loci

To leverage the shared genetic architecture between FGT polyps and related gynaecological disorders, multi-trait analysis of GWAS (MTAG) was performed, combining FGT polyps with endometrial cancer, ovarian cancer, endometriosis, and uterine fibroids. MTAG identified 47 genome-wide significant loci for FGT polyp risk (Figure 5), 26 of which were not detected in the primary meta-analysis, bringing the total to 52 risk loci across both analyses (**Supplementary Table 11**). MTAG genetic correlations ranged from r_g_ = 0.44 to 0.90 (**Supplementary Table 12**), with the strongest correlation with the FGT polyps meta-analysis (r_g_ = 0.90), confirming that MTAG predominantly captured FGT polyp signal. The multi-trait analysis also replicated 16 of 21 previously published polyp loci^7,9^. Six of the new MTAG loci mapped to genes prioritised by either TWAS or MAGMA, including rs28689909 (near estrogen regulator gene *GREB1*) and rs114088263 (near established oncogene *HMGA1*) ^20,21^.

**Figure 5.**
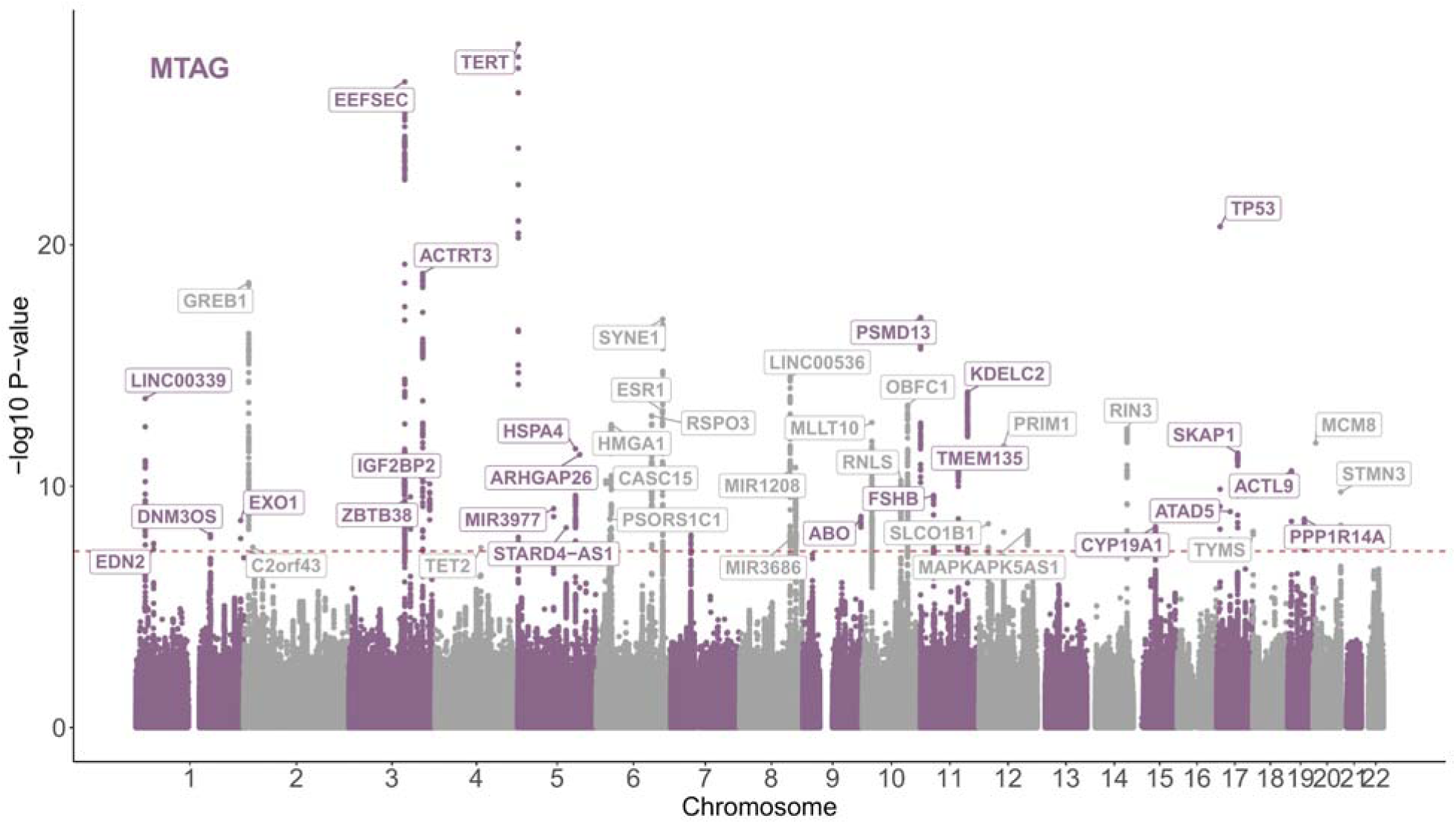
Multi-trait analysis (MTAG) identifies additional FGT polyp risk loci. Manhattan plot of MTAG results combining FGT polyps with genetically correlated gynaecological disorders (endometrial cancer, ovarian cancer, endometriosis, and uterine fibroids). Genome-wide significant loci (P < 5x10^-8^) are annotated with the nearest gene.

### Directional consistency in an independent replication cohort

We sought to replicate our findings in the All of Us cohort (3,812 cases, 130,368 controls). Reduced power in this smaller cohort precluded genome-wide significant replication; 9 nominally significant loci (P < 5×10) were identified, none overlapping meta-analysis or MTAG loci (**Supplementary Table 13).** Nevertheless, inverse-variance-weighted regression demonstrated significant directional concordance for the 26 lead meta-analysis SNPs (β = 0.62, 95% CIs = 0.42–0.83; **Supplementary Figure 5**), increasing to IVW = 0.95 (0.54–1.36) when including MTAG lead SNPs (**Supplementary Figure 6**). Of the 52 total risk loci, 39 (75%) showed a consistent direction of effect, including 18 where confidence intervals overlapped point estimates of the replication set (**Supplementary Figures 7** & **8**, **Supplementary Table 11**). The global genetic correlation between our meta-analysis and All of Us was moderate (r_g_ = 0.5, 95% CIs = 0.19–0.80), though LDSC accuracy is likely diminished given the absence of genome-wide significant signals in the replication set ^22^.

## Discussion

This study represents the largest genetic analysis of FGT polyps to date, identifying 26 independent risk loci from a meta-analysis of four European cohorts (48,400 cases and 477,134 controls), 12 of which were previously unreported. Fine-mapping revealed four missense variants, three predicted to be damaging, highlighting potential causal risk genes. By combining findings from gene-based testing, a previous FGT polyp GWAS analysis, and integration of functional genomic data at GWAS risk loci, we prioritised 193 candidate risk genes. Enrichment analysis of these genes revealed three dominant biological mechanisms: regulation of DNA replication machinery and genome maintenance, obesity-related drivers, and estrogen signalling.

Beyond specific loci, we defined the broader genetic architecture of FGT polyps through genetic correlation with other gynaecological diseases; and Mendelian randomisation revealed bidirectional causal relationships with endometrial cancer, endometriosis and uterine fibroids. Finally, leveraging the shared genetic architecture across four comorbid gynaecological disorders identified a further 26 loci via multi-trait analysis. In total, we identified 52 risk loci (including 36 that had not been previously reported ^7,9^), with 39 (75%) showing a consistent direction of effect in an independent replication cohort.

Convergent evidence from GWAS hits, fine-mapping, and pathway enrichment implicates defective DNA replication and genome maintenance as a core mechanism in FGT polyp pathogenesis. A primary locus maps to *TERT*, a key regulator of telomere maintenance ^23^, suggesting that sustained replicative potential is central to polyp susceptibility. This theme is reinforced by fine-mapping, which identified a high-confidence missense variant (PIP > 0.8) in *PRIM1*, which is essential for synthesising RNA primers during DNA replication; biallelic loss-of-function variants cause replication stress and profound growth defects ^24^. Given the cyclic, hormone-driven proliferation characteristic of the endometrium, even subtle variation in this machinery could plausibly impair fidelity in rapidly dividing epithelial cells. A missense variant was also identified in *ATM*, a master regulator of DNA damage response ^25^, though computational predictions suggest this variant is benign.

Consistent with these findings, gene set enrichment identified DNA replication as the most prominent biological process among candidate genes. Beyond *PRIM1*, this highlighted other functionally interconnected components of the replication machinery: CDT1, which loads the helicase complex (includes the protein encoded by the candidate *MCM6* gene) onto DNA replication origins ^26^; STN1, which facilitates replication through difficult regions and promotes fork restart ^27^; and chromatin factor HMGA1, which regulates expression of replication-dependent histone genes required for chromatin assembly during DNA synthesis ^28^. *HMGA1* represents a compelling example of germline-somatic convergence: initially sub-threshold in gene-based analysis, it was validated as genome-wide significant in the multi-trait analysis, and chromosomal rearrangements affecting HMGA1 are reported in up to 74% of endometrial polyps ^14,29^, indicating that both inherited variation and acquired somatic alterations converge on this replication-regulating chromatin factor. Collectively, these findings suggest germline variation in replication fidelity creates a proliferative state that increases polyp risk ^7,9^ ^30^ ^31,32^.

Polyps have been clinically associated with conditions of estrogen exposure, including hormone replacement therapy, tamoxifen use, and obesity ^33^. Gene set enrichment directly supported these associations, with candidate genes enriched for hormone-related gynaecological disorders, obesity-related traits, and estrogen responses. Key estrogen signalling genes included *ESR1*, encoding estrogen receptor alpha, the most prominent estrogen receptor in the endometrium ^34^; *GREB1*, a direct transcriptional target of the estrogen receptor that regulates cell proliferation in hormone-responsive tissues ^35,36^, and has been implicated in other estrogen-related gynaecological disorders ^30,37^; *CYP19A1* encoding aromatase, the rate-limiting enzyme converting androgens to estrogens. Notably, HMGA1 enhances estrogen receptor binding to its response element ^13^, further linking replication machinery to hormonal signalling. Together, these findings support a model wherein inherited FGT polyps susceptibility spans the estrogen signalling axis, from hormone biosynthesis to receptor-mediated transcriptional regulation.

This hormonal aetiology is further supported by the capacity of adipose tissue to synthesise estrone via peripheral aromatisation ^38^, underscored by our identification of *CYP19A1*. Consistent with this pathway, multiple obesity-associated candidate genes were identified, including *RSPO3*, a Wnt signalling modulator influencing body fat distribution and adipose cell biology ^39,40^; *PLCE1*, associated with body fat distribution ^41^; and *IGF2BP2*, a regulator of hepatic fatty acid oxidation and triglyceride accumulation ^42^.

The central role of estrogen signalling is further underscored by the strong genetic correlations observed between FGT polyps and other hormone-related gynaecological disorders (**Figure 4**). Most notably, endometriosis and uterine fibroids, both characterised by aberrant cellular growth, suggest a substantial shared genetic architecture across a spectrum of estrogen-responsive conditions. We leveraged this overlap via MTAG to identify 26 additional risk loci, capturing variants that would otherwise require larger sample sizes to detect, and elevating six biologically relevant sub-threshold signals from the base meta-analysis (including *HMGA1* and *GREB1)* to genome-wide significance, suggesting the additional MTAG loci identified represent *bona fide* polyp risk variants. Conversely, the absence of genetic correlation with cervical cancer reflects its distinct non-hormonal, virally driven aetiology ^43^.

The bidirectional causal relationships with endometrial cancer, endometriosis, and uterine fibroids are particularly striking; they suggest these associations reflect common underlying biological pathways rather than chance co-occurrence. While sample overlap may partially contribute, the persistent associations after correction for endometrial cancer support genuine shared genetic architecture. FGT polyps showed no causal effect on breast cancer; however, the reverse association — breast cancer liability increasing polyp risk — likely reflects pleiotropic effects of estrogen signalling variants on endometrial proliferation^17,18^.

These genetic findings are supported by substantial clinical comorbidity in the UK Biobank, where polyp cases had markedly elevated odds of uterine fibroids (OR = 5.16) and higher prevalence of endometriosis (5.96% in polyps cases vs 1.78% controls). Although detection bias likely contributes to this clinical overlap, the robust genetic correlations imply that shared biological susceptibility also plays a role, positioning FGT polyps within a broader polygenic landscape of gynaecological growth disorders.

Our study has several limitations. First, average FGT polyp prevalence across cohorts was 7.9%, below population estimates of 8-50% ^1–3^, suggesting undiagnosed cases among controls — particularly in All of Us (2.9% prevalence). This may contribute to inter-cohort genetic correlation heterogeneity (**Supplementary Figure 1**). SNP effect heterogeneity (I²) for lead SNPs ranged from 0–75 (mean = 32.71; Supplementary Table 4), suggesting some loci may be cohort-driven, though no lead SNP showed significant heterogeneity after multiple testing correction. Differences in screening uptake across countries may also contribute to prevalence differences between cohorts ^44^. Second, all GWAS cohorts were of European ancestry, limiting generalisability. Third, the absence of Finnish- or Estonian-specific LD reference panels reduced fine-mapping accuracy at some loci. Additionally, our gene prioritisation strategy favoured sensitivity, aggregating candidates from multiple mapping approaches, but limits specificity, potentially capturing non-causal ’bystander’ genes in complex LD regions. Furthermore, reliance on HiChIP data from an immortalised endometrial cell line may not fully capture the distinct regulatory landscape of primary polyp tissue. Finally, comparison with a bioavailable estrogen GWAS was not possible; while a GWAS for total estradiol exist^45^, cycle-phase variability and assay sensitivity limitations restrict their utility in this context.

In conclusion, this large-scale meta-analysis identifies 52 risk loci for FGT polyps — 26 from primary meta-analysis and 26 from multi-trait analysis — of which 36 were novel and 39 (75%) replicated directionally in an independent cohort. Integration of gene-based and functional genomic approaches prioritised 193 candidate genes, with pathway enrichment revealing a convergent mechanism whereby germline variation in replication machinery compromises genomic stability in the context of hormone-driven endometrial proliferation. This susceptibility is further modulated by metabolic drivers of estrogen biosynthesis, underscored by the identification of specific adiposity-related loci and *CYP19A1*. Collectively, these findings position polyps not as isolated lesions but as manifestations of a broader gynaecological proliferative syndrome shared with endometriosis and uterine fibroids. This is further supported by the strong genetic correlations observed between polyps and other gynaecological disorders, with Mendelian randomisation revealing bidirectional causal relationships with endometriosis, uterine fibroids and endometrial cancer risk. These findings provide a foundation for genetic risk stratification across diverse ancestries and suggest that therapeutic strategies targeting the estrogen-replication axis or metabolic health could offer non-surgical avenues for prevention and management.

## Methods

### FGT polyp GWAS cohorts

Data from four cohorts were included in the main GWAS meta-analysis: UK Biobank, FinnGen (R11) ^46^, Estonian Biobank and the Michigan Genomics Initiative (Freeze 3) ^47^, totalling 48,400 cases and 477,134 controls. An independent replication GWAS was performed in 3,812 cases and 130,368 controls in the All of Us cohort. Cohort descriptions, recruitment, specific genotyping, imputation and GWAS analysis details are provided in the Supplementary Note and **Supplementary Table 14**. Briefly, cases were identified via ICD-8, 9, or 10 codes from electronic health records. GWAS were performed in homogeneous European ancestral populations using REGENIE or SAIGE, with one monozygotic twin per pair excluded. Standard participant- and genotype-level QC was applied (**Supplementary Table 14**).

### GWAS Meta-analysis

All datasets were harmonised to GRCh38 using UCSC LiftOver ^48^. SNPs with imputation quality INFO < 0.3, MAF < 0.01, or extreme effects (OR > 5 or < 0.2) were excluded. SNP effect and standard error estimates were meta-analysed across four cohorts using a fixed-effects inverse-variance weighted approach in METAL v.3 ^49^(**Figure 1**; **Supplementary Table 14**), retaining only SNPs present in at least two contributing datasets.

### Lead variant and credible set identification

Independent lead SNPs at genome-wide significant loci (P < 5×10) were identified using the ’clump’ function in PLINK v.1.9 (r² < 0.001, distance > 1000 kb) with a UK Biobank LD reference panel ^50^. Lead SNPs were then used to define credible sets of candidate causal variants (CCVs) via cross-population fine-mapping with SuSiEx, which assigns posterior inclusion probabilities (PIPs) to each variant ^12^.

### Candidate gene prioritisation and gene-set enrichment

Three approaches were used to prioritise candidate target genes of FGT polyp-associated SNPs identified by GWAS meta-analysis: Multi-marker Analysis of GenoMic Annotation (MAGMA), Transcriptome wide association study (TWAS) and interrogation of HiChIP chromatin interaction data from endometrial cells.

MAGMA aggregates SNP-level signals across genes and was run using MAGMA v.1.08 ^51^ with the European 1000 Genomes Project v3 reference panel and proximity-based SNP-to-gene mapping. The significance threshold was P < 2.5×10 (Bonferroni correction for 20,174 protein-coding genes).

TWAS was performed on meta-analysis summary statistics using FUSION v.1 ^52^, using the default parameters with precomputed eQTL models from GTEx v8 (ovary, vagina, whole blood, EBV-transformed lymphocytes) and cross-tissue sCCA models ^15^. The *MHC* region on chromosome 6 was removed post-analysis.

E6E7 hTERT immortalised endometrial epithelial cells underwent H3K27Ac HiChIP analysis as previously described ^53^. Briefly, a HiC+ Kit (Arima Genomics) generated global chromatin contact libraries, and fragmented chromatin was captured using an antibody raised against H3K27Ac (Abcam, EP16602). After sample clean-up and tagmentation, PCR sequencing libraries were generated. Two independent HiChIP sequencing libraries were analysed by 2×150bp sequencing using an Illumina NextSeq550 (QIMRB Sequencing Facility, Brisbane, Australia). HiChIP reads were mapped to the human reference genome (build hg38) using HiC-Pro v2.11.4 ^54^, using default settings to remove duplicate reads and filter for valid interactions.

Statistically significant contacts were identified using ‘peak-to-all’ interactions FitHiChIP v8.1.0 (FDR < 5%) using the inferred H3K37Ac peaks called from the one-dimensional HiChIP data and bin size of 5 kb. Distance thresholds were set at 20 kb – 2Mb for interactions and bias correction was performed using coverage-specific bias. Using the UCSC Genome Browser (https://genome.ucsc.edu/; hg38 accessed November 2025) ^55^, we intersected anchors of H3K27Ac HiChIP chromatin loops with 636 GWAS meta-analysis CCVs from SuSiEx and promoter regions (within 3kb of transcription start sites) to identify candidate target genes at FGT polyp risk loci.

Candidate genes from all three approaches, together with those from Pathare et al., were submitted to ToppFun ^16^, using default settings, including a Benjamini-Hochberg False Discovery Rate (FDR) correction to retain only those terms with an adjusted p-value below 0.05.

### Genetic correlation with gynaecological traits

SNP-based heritability and genetic correlations (rg) between FGT polyps and six gynaecological traits: endometrial cancer ^56^, uterine fibroids ^57^, polycystic ovarian syndrome (PCOS) ^58^, endometriosis ^37^, cervical cancer ^59,60^, ovarian cancer ^61^ were estimated using Linkage Disequilibrium score regression (LDSC). A Bonferroni corrected p value < 0.0083, correcting for 6 traits tested, was applied to mark significance.

### Mendelian Randomisation

Bidirectional MR was performed between FGT polyps and the above traits, plus breast cancer ^17,18^, using lead FGT polyp meta-analysis SNPs (P < 5×10) as instrumental variables in an inverse-variance weighted (IVW) regression with the SNPs estimated effect on each disorder within the MR-Base R package ^62,63^. Instrument strength was assessed via F-statistic approximation (F ≈ (β/SE)²; strong IVs defined as mean F > 10). MR-PRESSO and a HEIDI outlier test were used to remove SNPs causing significant levels of heterogeneity, violating the pleiotropy assumption ^64^. The MR-Egger intercept was used to assess residual pleiotropy, where heterogeneous SNPs were removed until the intercept was not significantly different from zero^63,65^.

### Multi-trait Analysis of GWAS

For traits with a bi-variate global r_g_ > 0.25 and P value < 0.0083, we performed MTAG with FGT polyps as the focal trait ^66^. Input GWAS were lifted over to hg38 and filtered (MAF > 1%, INFO > 0.3) prior to running MTAG with default settings.

### Replication

The All of Us GWAS was performed on the All of Us research workbench and clumped using PLINK v.1.9 (r² < 0.001, P < 5×10, distance > 1000 kb); full details are in the Supplementary Notes. Lead SNPs from the meta-analysis and MTAG (or proxies with r² > 0.98) were extracted and effect estimates compared in R v4.5.0. A locus was considered replicated if the direction of effect matched.

### Ethics

UKBB participants gave informed consent provided under the ethical approval of the Northwest Multi-centre Research Ethics Committee. This project used data from the UKBB under application number 25331. All Estonian Biobank participants have signed a broad informed consent for using their data in research, and the study was carried out under ethical approval from the Estonian Committee on Bioethics and Human Research (Estonian Ministry of Social Affairs). FinnGen Participants gave informed consent under the ethical approval of the Coordinating Ethics Committee of the Hospital District of Helsinki and Uusimaa. For this study, we downloaded the publicly available summary statistics from the FinnGen data portal Release 11. Participants provided informed consent under ethical approval from The University of Michigan Medical School Institutional Review Board. We accessed GWAS summary statistics of the MGI data for ‘freeze 3’. All of Us participant data was collected with informed consent under the ethical approval and ongoing oversight of the All of Us Institutional Review Board. We accessed and analysed ‘controlled’ tier short read whole genome sequencing data through the online All of Us researcher workbench. This study was conducted under the ethical approval of the QIMR Berghofer ethics committee (P1051).

## Data Availability

GWAS summary statistics will be made publically available on Zenodo at time of publication.

## Acknowledgements

The authors acknowledge research participants and research staff of the UK Biobank, Estonian Biobank, FinnGen, The Michigan Genomics Initiative, and the All of Us cohort. We thank the GWAS Catalog for storing and facilitating access to publicly available GWAS summary statistics. We thank the QIMR Berghofer Genome Informatics team for maintaining the high-performance computing cluster and providing computational advice.

This work was supported by a National Health and Medical Research Council (NHMRC) of Australia Investigator Grants (APP1173170 and #ID 2041819) awarded to T.A.O’M, and an NHMRC Project Grant (APP1158083) awarded to T.A.O’M and D.M.G. K.B. is supported by a QIMR Berghofer Cancer Genetic Susceptibility PhD Scholarship and a University of Queensland Research Training Scholarship. T.L and J.D was supported by the Estonian Research Council grant PSG776. S.F was supported by the QIMR Berghofer Vocational Research Scholarship. The funders had no role in study design, data collection and analysis, interpretation of results, or manuscript preparation.

## Author Contributions

N.I. wrote the main manuscript, analysed data, prepared all main figures, and led the interpretation of results. K.B., S.F., E.M., D.C., J.D. and T.L. analysed data, contributed to the interpretation of results and generated some supplementary figures and tables. S.Z., S.P., and T.L. contributed to the collected cohort data. D.M.G. and T.A.O.M analysed data, led the interpretation of results, and supervised the project. All authors read and contributed to the drafting of the final manuscript.

## Notes

### Competing Interest Statement

The authors have declared no competing interest.

### Author Declarations

UKBB participants gave informed consent provided under the ethical approval of the Northwest Multi-centre Research Ethics Committee. This project used data from the UKBB under application number 25331. All Estonian Biobank participants have signed a broad informed consent for using their data in research, and the study was carried out under ethical approval from the Estonian Committee on Bioethics and Human Research (Estonian Ministry of Social Affairs). FinnGen Participants gave informed consent under the ethical approval of the Coordinating Ethics Committee of the Hospital District of Helsinki and Uusimaa. For this study, we downloaded the publicly available summary statistics from the FinnGen data portal Release 11. Participants provided informed consent under ethical approval from The University of Michigan Medical School Institutional Review Board. We accessed GWAS summary statistics of the MGI data for freeze 3. All of Us participant data was collected with informed consent under the ethical approval and ongoing oversight of the All of Us Institutional Review Board. We accessed and analysed controlled tier short read whole genome sequencing data through the online All of Us researcher workbench. This study was conducted under the ethical approval of the QIMR Berghofer ethics committee (P1051).

