## Supplementary Figures for "Genetic analysis of female genital tract polyps implicates genome stability, estrogen signalling and shared susceptibility with proliferative gynaecological disorders"

### Cohorts

Before the GWAS analysis was performed, participants with extreme heterozygosity, participants and Single Nucleotide Polymorphisms (SNPs) with a high level of missingness, SNPs that were poorly imputed, SNPs significantly deviating from Hardy-Weinberg equilibrium, and SNPs with a minor allele frequency < 1% were removed. The P value threshold to mark genome-wide significance was set to P < 5 × 10-8. Specific methods for ancestral determination differ between cohorts, though all were filtered to a homogenous ancestral population. This resulted in 8,310,442 SNPs tested to provide broad whole genome coverage (chromosomes 1-23) across all 4 cohorts

#### UK Biobank

The first cohort analysed was the UK Biobank (UKBB), which is an open-access population-based prospective cohort study including 273,405 women from across the United Kingdom, aged 40–69 years at recruitment (between 2007 and 2010). UKBB participants gave informed consent provided under the ethical approval of the Northwest Multi-centre Research Ethics Committee [1]. We accessed participant's genetic and health-registry data (updated as of February 2024) under application 25331. Before analysis, participants were filtered to a homogenous European population by excluding samples where the first two genetic Principal Components (PCs) fell outside (greater than 3 standard deviations away from) the center of the largest self-report 'white-British' PCs cluster from all UKBB samples.

#### FinnGen (r11)

For the second cohort, we accessed release 11 of the FinnGen cohort, including 282,064 women from Finland, ages ranging from 18 to over 70 years. FinnGen participants were recruited from 2017 - 2024. Participants gave informed consent under the ethical approval of the Coordinating Ethics Committee of the Hospital District of Helsinki and Uusimaa [2]. PC analysis was used to filter participants to a homogenous ancestral population using a Finnish-specific subset of the 1000 Genomes Project as a reference population. One of each second-degree relative, along with participants with a mismatch between genotypic and phenotypic sex were also removed [3].

##### Estonian Biobank

The Estonian Biobank (EstBB) is an Estonian-based cohort, including ~135,000 women, with a mean age of 49.2 years at the time of recruitment (2000-2024). Recruitment of EstBB participants was performed with informed participant consent under the ethical approval of the Estonian Committee on Bioethics and Human Research (Estonian Ministry of Social Affairs) [4]. All individuals included in this study are of European ancestry, as inferred from genetic grouping. One sibling from each twin/sibling pair within the cohort were removed, along with samples with sex mismatch.

##### Michigan Genomics Initiative

The Michigan Genomics Initiative (MGI) consists of ~37,000 women, with a mean age of 57 at the time of recruitment (2015-current). Participants provided informed consent under ethical approval from The University of Michigan Medical School Institutional Review Board (IRB) [5]. We accessed 'freeze 3', which involved participants being filtered to a homogenous European ancestry using the first two PCs and a Human Genome Diversity Project reference population. Participants with a sex mismatch between genotype and phenotype were removed.

##### All of Us

To replicate the findings of the resulting GWAS meta-analysis and the multi-trait analysis, we used the All of Us cohort (**Supplementary Table 1**). All of Us consists of 134,109 women of European descent. Recruitment started in 2017 and was conducted with informed consent under the ethical approval and ongoing oversight of the All of Us Institutional Review Board [6]. We accessed and analysed ‘controlled’ tier short read whole genome sequencing data through the online All of Us researcher workbench. 3812 FGT polyp cases were selected using ICD9, 10 and SNOMED codes. Samples were filtered to a homogenous European sample using PC analysis compared to 1000 genomes and Human Genome Diversity Project reference sample [7].

**Supplementary Figure 1: Linkage Disequilibrium score regression estimates between the cohort-level GWAS used in the GWAS meta-analysis, the GWAS meta-analysis and the replication GWAS.**

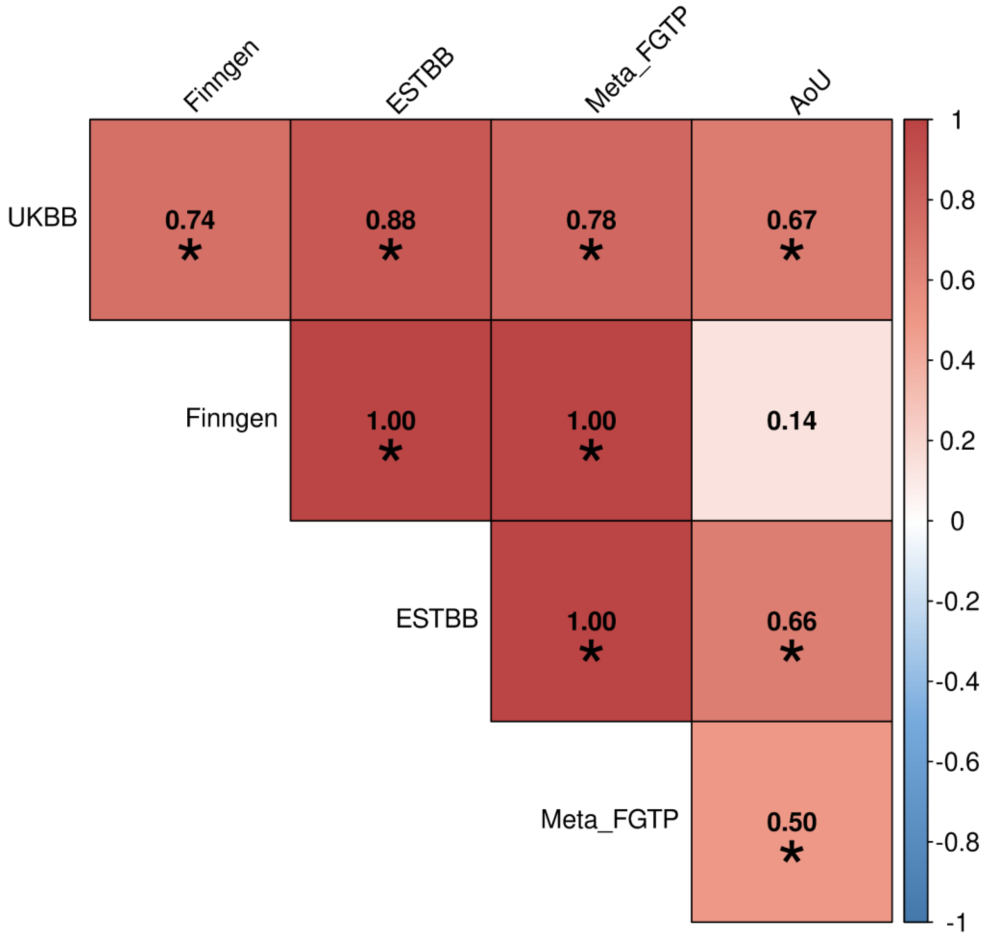


***Supplementary Figure 1:*** *A heatmap showing the genetic correlation (rg) between the GWAS meta-analysis input GWASs (UK Biobank [UKBB], FinnGen and Estonian Biobank [EstBB]). The plot also includes the replication sets All of Us (AoU). The main meta-analysis (Meta_FGTP) is also included. The Michigan Genomics Initiative is not included in the plot as the sample size was too low to calculate a meaningful SNP-based heritability and rg estimate.*

**Supplementary Figure 2: Venn diagram showing all 194 genes prioritised by 1 of 4 analyses for female genital tract polyps**

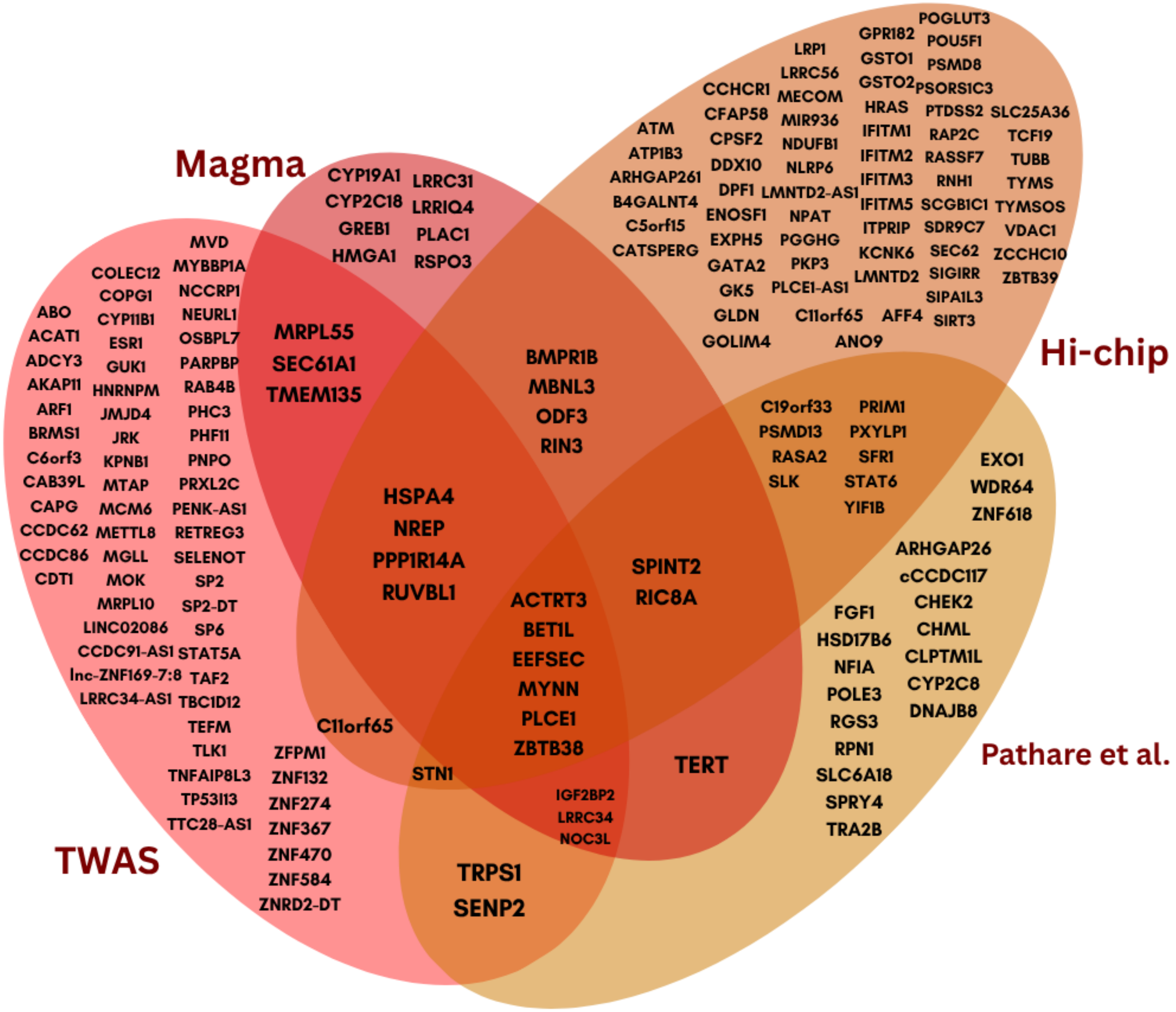


***Supplementary Figure 2***: *Venn diagram showing all 193 genes prioritised by 1 of 3 analyses performed in this paper for female genital tract polyps, plus those reported from the MAGMA analysis in Pathare et al., 2025*

**Supplementary Figure 3: Forest plot of the Mendelian Randomisation estimating the effect of Female genital tract polyps on eight traits**

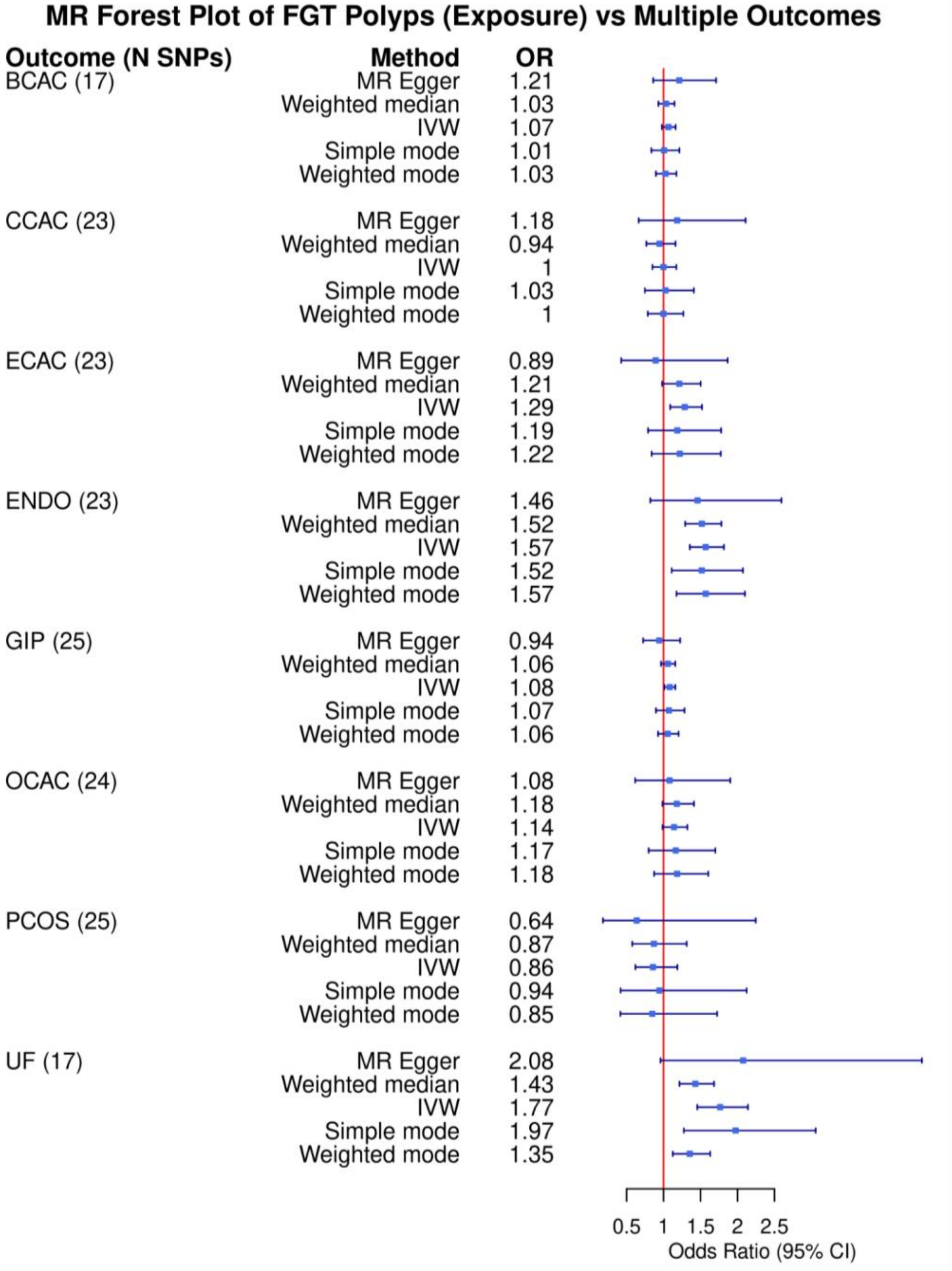


***Supplementary Figure 3**: A forest plot showing the odds ratios and 95% confidence intervals from 5 different regression methods (inverse variance weighted [IVW], plus 4 sensitivity analyses). Each model indicates a change in odds of a disease given an increased genetic liability of female genital tract polyps. The eight traits include: Breast cancer (BCAC), Cervical cancer (CCAC), Endometrial cancer (ECAC), Endometriosis (ENDO), Gastro-intestinal polyps (GIP), Ovarian cancer (OCAC), Polycystic ovarian syndrome (PCOS), and Uterine fibroids (UF). Also shown is the number of single nucleotide polymorphisms (SNPs) to predict FGT polyps genetic liability in*

*each model; this number differs between traits, as SNPs will be removed if they have an independent effect on the outcome disease, or the SNP has no available effect estimates on the outcome.*

**Supplementary Figure 4: Forest plot of the Mendelian Randomisation estimating the effect of eight traits on female genital tract polyp risk**

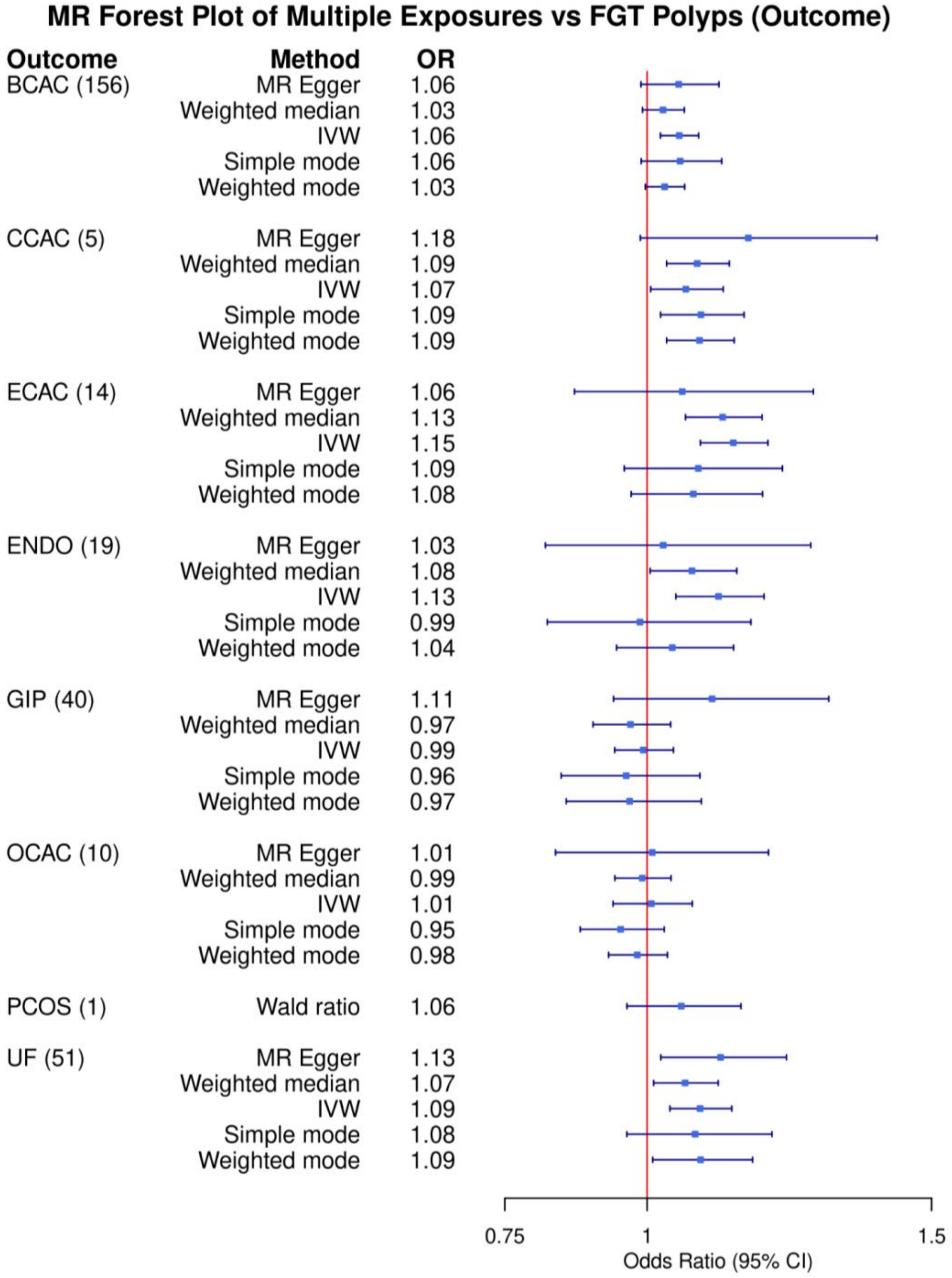


***Supplementary Figure 4****: A forest plot showing the odds ratios and 95% confidence intervals from 5 different regression methods (inverse variance weighted [IVW], plus 4 sensitivity analyses). Each model indicates a change in the odds of female genital tract polyps given an increased genetic liability of a disease. The eight traits include: Breast cancer (BCAC), Cervical cancer (CCAC), Endometrial cancer (ECAC), Endometriosis (ENDO), Gastro-intestinal polyps (GIP), Ovarian cancer (OCAC), Polycystic ovarian syndrome (PCOS), and Uterine fibroids (UF).*

**Supplementary Figure 5: Inverse variance weighted regression of the effect estimate of the lead loci of the GWAS meta-analysis and the replication dataset.**

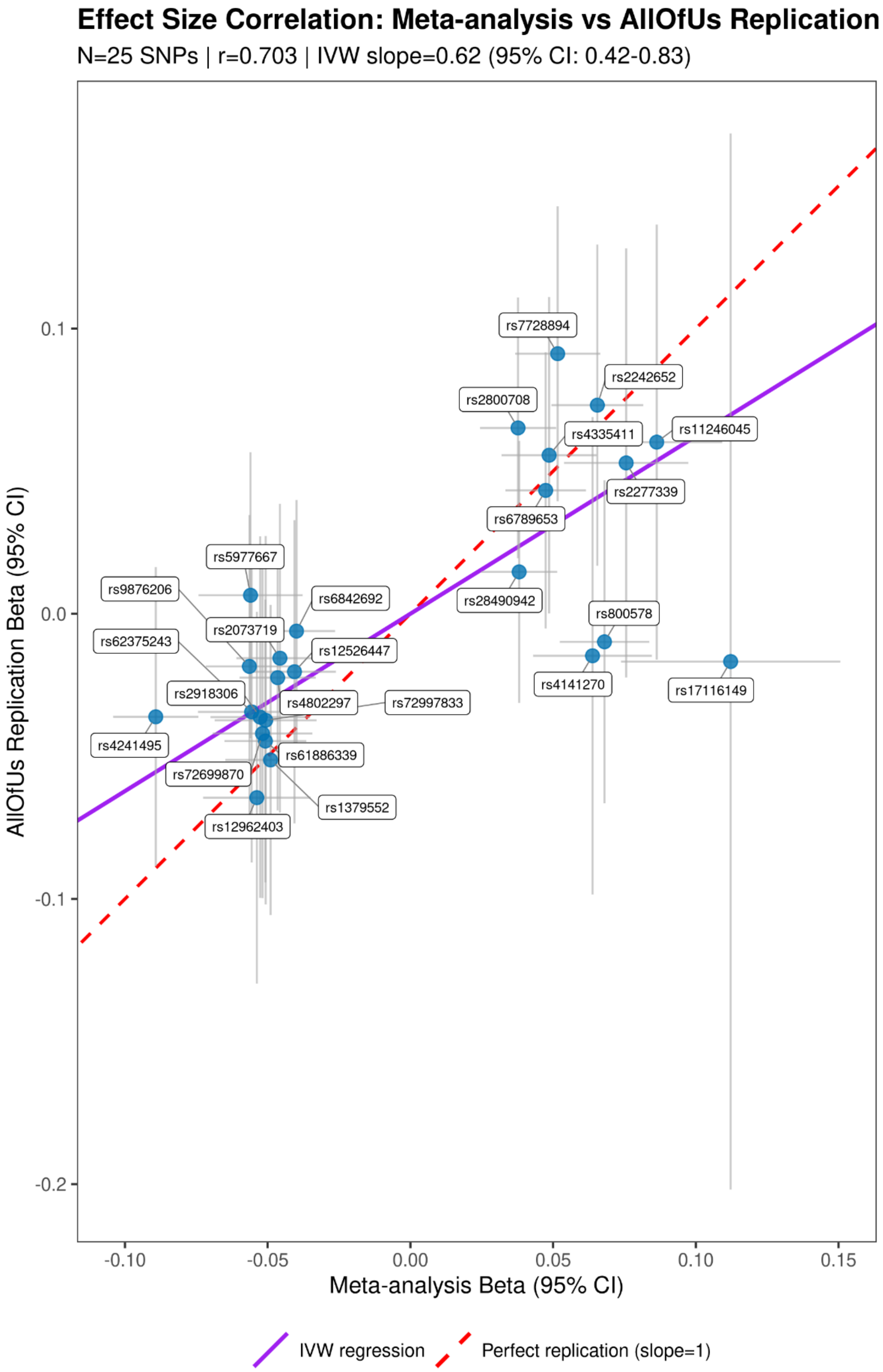


***Supplementary Figure 5:*** *Scatter plot of effect estimates and standard errors of the 25 of the 26 lead SNPs for FGT polyps from the GWAS meta-analysis SNPs from the All of Us cohort. The*

*purple line represents inverse variance weighted regression, and the red dotted line is a perfect correlation.*

**Supplementary Figure 6: Inverse variance weighted regression of the effect estimate of the lead loci of the GWAS meta-analysis + MTAG and the replication dataset.**

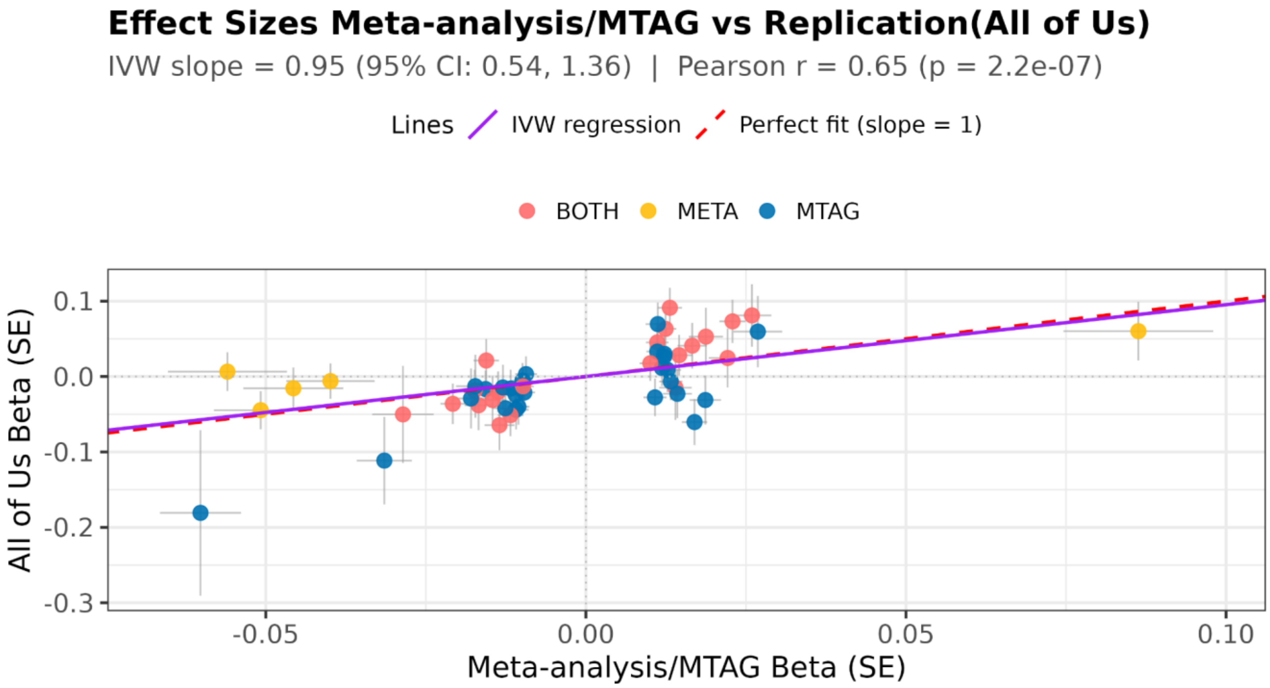


***Supplementary Figure 6:*** *Scatter plot of effect estimates and standard errors of the lead SNPs for the GWAS meta-analysis and MTAG of FGT polyps compared to the SNPs' estimated effects within the All of Us cohort. The purple line represents inverse variance weighted regression, and the red dotted line is a perfect correlation.*

**Supplementary Figure 7: Forest plot comparing effect estimates of the GWAS meta-analysis and the replication GWAS of FGT polyps for 25 significant loci**

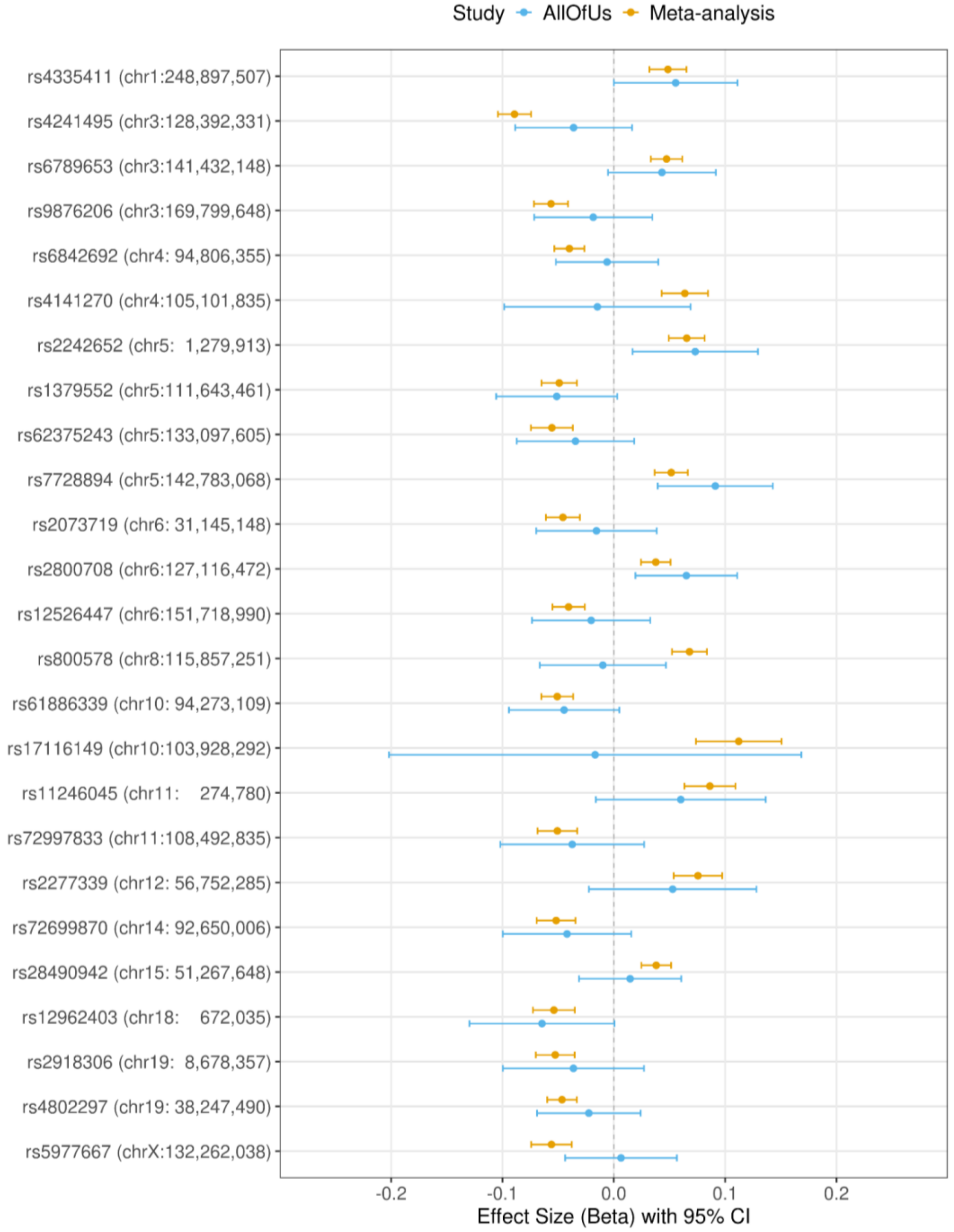


***Supplementary Figure 7:*** *A forest plot showing 25 of the 26 lead GWAS meta-analysis effect estimates and 95% confidence intervals for the GWAS meta-analysis (orange) and the replication data set (blue). One SNP rs674311 was not present in the replication data set, nor was a suitable LD matching proxy.*

**Supplementary Figure 8: Forest plot comparing effect estimates of the MTAG and the replication GWAS of FGT polyps for 46 significant loci**

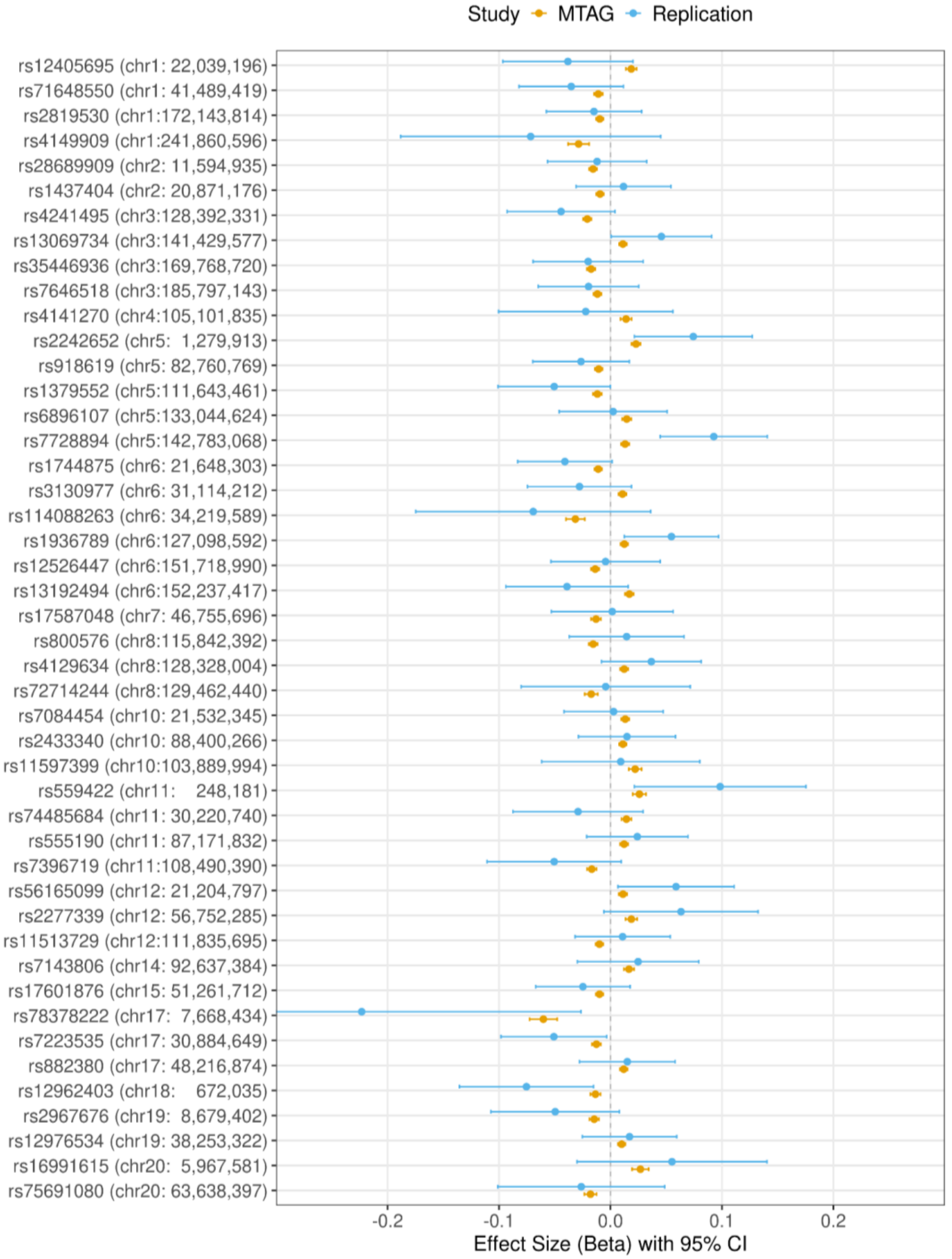


***Supplementary Figure 8:*** *A forest plot showing 46 of the 47 lead GWAS multi-trait analysis of GWAS effect estimates and 95% confidence intervals for the MTAG (orange) and the replication data set (blue). One SNP,* rs635634, *was not present in the replication data set, nor was there a suitable LD matching proxy.*
